# Evaluating the Clinical Impact of CYP2C19 and CYP2D6 on Amitriptyline Outcomes in a Real-World Chronic Pain Cohort

**DOI:** 10.64898/2026.05.28.26354228

**Authors:** Bade Uckac, Santiago Díaz-Torres, Zuriel Ceja, Natalia S. Ogonowski, Penelope A. Lind, Dale R. Nyholt, Nicholas G. Martin, Sarah E. Medland, Miguel E. Rentería, Giovanni E. Ferreira

**Affiliations:** Brain & Mental Health Program, QIMR Berghofer Medical Research Institute, Brisbane, QLD, Australia; School of Biomedical Sciences, Faculty of Health, Queensland University of Technology, Brisbane, QLD, Australia; Population Health Program, QIMR Berghofer Medical Research Institute, Brisbane, QLD, Australia; School of Biomedical Sciences, Faculty of Health, Medicine and Behavioural Sciences, The University of Queensland, Brisbane, QLD, Australia; School of Psychology, The University of Queensland, Brisbane, QLD, Australia; School of Psychology and Counselling, Queensland University of Technology; Institute for Musculoskeletal Health, Sydney Local Health District, Sydney, NSW, Australia; Sydney School of Public Health, The University of Sydney, Sydney, NSW, Australia

**Author notes:** Correspondence: Miguel E. Rentería and Giovanni E. Ferreira.

**Keywords:** pharmacogenetics, chronic pain, depression, CYP2C19, CYP2D6, amitriptyline, translational medicine

## Abstract

**Background:** Amitriptyline is widely prescribed for chronic pain, yet treatment response and tolerability vary substantially. Genetic variation in CYP2C19 and CYP2D6 influences amitriptyline metabolism, but evidence linking pharmacogene status to clinical outcomes in chronic pain populations remains limited. Importantly, amitriptyline is typically prescribed at lower doses for chronic pain (10–50 mg/day) than for depression (75–150 mg/day), which may attenuate observable pharmacogenomic effects. We investigated whether *CYP2C19* and *CYP2D6* derived metaboliser phenotypes were associated with self-reported effectiveness and tolerability of amitriptyline.

**Methods:** We conducted a cross-sectional analysis within the Australian Genetics of Depression Study. Participants reporting chronic pain, amitriptyline use, treatment response data, and genotype data were included (n=1146). Pharmacogene variants were annotated using PharmCAT, and plausible diplotypes were harmonised using averaged gene-activity scores and a CPIC-guided framework to assign final metaboliser phenotypes. Associations with effectiveness and tolerability were examined using multinomial and binary logistic regression models adjusted for age and sex.

**Results:** Reported effectiveness was generally low: 46.0% reported amitriptyline worked not at all well, 36.6% moderately well, and 17.4% very well. Among participants with tolerability data, 53% discontinued treatment because of side effects. *CYP2D6* phenotype was not associated with effectiveness or discontinuation. *CYP2C19* intermediate metabolisers showed nominally reduced odds of discontinuation, although this did not survive multiple-testing correction.

**Conclusions:** *CYP2C19* and *CYP2D6* derived phenotypes were not strongly associated with amitriptyline effectiveness or tolerability in this chronic pain cohort with comorbid mental health disorders, suggesting that low-dose prescribing and titration-based management may attenuate pharmacogenomic effects in routine care.

## INTRODUCTION

Chronic pain (lasting more than three months) remains a clinical challenge because it is biologically and psychologically complex, varies widely between individuals, and often persists despite standard treatments. Its mechanisms frequently extend beyond the original injury, involving central sensitisation and neuroimmune changes, and it commonly co-occurs with depression, anxiety, sleep problems, and functional impairment.^1, 2, 3^ In Australia, chronic pain affects approximately one in five adults aged 45 and over (around 3.37 million people) and this number is projected to exceed 5 million by 2050, with total costs expected to surpass $215 billion by then, according to the latest research ^4^.

Antidepressants are frequently prescribed off-label for chronic pain conditions ^5,6^, including lower back pain and osteoarthritis.^7^ Tricyclic antidepressants (TCAs), particularly amitriptyline, are among the most commonly prescribed antidepressants for chronic pain worldwide.^8^ In Australia, amitriptyline is the leading antidepressant for patients using opioids for chronic pain (17%) and for those with lower back pain in the workers’ compensation system (55%).^9^ Its use related to pain is also high in other countries, such as the Netherlands ^10^ and the UK.

Although amitriptyline has demonstrated efficacy in neuropathic pain, irritable bowel syndrome, and chronic tension-type headache, evidence for its effectiveness in other chronic pain conditions, such as lower back pain and fibromyalgia, remains inconsistent.^11^ Despite this uncertainty, amitriptyline continues to be widely prescribed, accompanied by high rates of treatment discontinuation. Understanding sources of inter individual variability may help clarify who is most likely to tolerate and persist with treatment in real-world chronic pain populations.^12^

Critically, amitriptyline is typically prescribed for chronic pain at substantially lower doses than for depression. Whereas antidepressant regimens commonly employ 75–150 mg/day, chronic pain guidelines generally recommend initiation at 10–25 mg/day with gradual titration, often remaining below 50 mg/day ^13^. At these lower doses, plasma concentrations of amitriptyline and its active metabolite nortriptyline may remain below the threshold at which inter-individual metabolic differences are clinically significant, potentially attenuating observable pharmacogenomic effects on treatment outcomes. This dose-effect consideration is essential for interpreting pharmacogenetic associations in chronic pain populations.

Adverse effects are common with amitriptyline and frequently limit tolerability, with chronic pain studies reporting anticholinergic symptoms, sedation, dizziness, and occasional cardiovascular events.^14^ These effects may be amplified in *CYP2D6* poor metabolisers (PM), who exhibit reduced metabolic clearance and consequently higher concentrations of active metabolites, increasing the risk of dose-related adverse effects. Patients taking amitriptyline are over seven times more likely to experience an adverse drug response compared to placebo, an effect that appears to be dose-independent.^15^ Identifying biomarkers to guide targeting amitriptyline to patients most likely to benefit from, or be adversely affected by, treatment is highly desirable.

Amitriptyline is primarily metabolised by the cytochrome P450 enzymes *CYP2C19* and *CYP2D6*, which influence plasma concentrations of the parent drug and its active metabolites.^16^ These enzymes mediate metabolic pathways that contribute to interindividual variability in drug response and side-effect profiles by influencing plasma levels of amitriptyline and its metabolites.^17^ Such variation can affect both pain relief outcomes and the risk of adverse events. However, the magnitude of this effect may vary across patients’ medical histories or in the context of gene-environment interactions. ^18^

Genetic variation in *CYP2D6* and *CYP2C19* meaningfully affects the metabolism of amitriptyline and its active metabolites. Individuals with reduced metabolic activity (PM or intermediate metabolisers (IM)) typically exhibit higher plasma concentrations of parent drug and/or metabolites, which have been associated with a greater risk of adverse effects. These pharmacokinetic differences have been demonstrated in multiple therapeutic drug monitoring and pharmacokinetic studies.^16, 19^ Conversely, those with rapid (RM) or ultra-rapid (UM) metabolic activity may experience subtherapeutic exposure and reduced efficacy.^20^ Some large-scale studies have been conducted to support the clinical relevance of *CYP2C19* and *CYP2D6* status in predicting antidepressant response and tolerability.^21^ However, the majority of these studies examined antidepressant response broadly in psychiatric populations, rather than amitriptyline-specific outcomes in chronic pain.

Critically, no large-scale study has systematically examined amitriptyline-specific outcomes in people with chronic pain or formally evaluated *CYP2C19* × *CYP2D6* metaboliser interactions, despite their sequential and complementary roles in amitriptyline metabolism, where variation in one enzyme can modify the consequences of variation in the other. ^22^ Moreover, prior research on amitriptyline in chronic pain has predominantly focused on narrow secondary pain conditions, such as diabetic neuropathy ^23^, or has been limited by small sample sizes, restricting generalisability to chronic pain as a broader and primary phenotype.^24–26^ Although the Dutch Pharmacogenetics Working Group (DPWG) provides dosing recommendations supported by pharmacokinetic evidence, robust validation linking combined *CYP2C19* and *CYP2D6* variation to real-world treatment effectiveness and tolerability in chronic pain populations remains lacking. To address these gaps, we leveraged data from the Australian Genetics of Depression Study (AGDS) to examine amitriptyline-specific outcomes in individuals with chronic pain, using a scalable computational framework to translate genotype into ordered metaboliser phenotypes and to evaluate gene-specific and combined metabolic effects in a large, comorbid population.

## METHODS

### Study design

We conducted a cross-sectional study reported in accordance with the Strengthening the Reporting of Observational Studies in Epidemiology (STROBE) statement. The AGDS study was approved by the QIMR Berghofer Medical Research Institute Human Research Ethics and Safety Committee (Approval Number: P2118).

### Data sources

We used data from the AGDS cohort (n ≈ 20,000), a large cohort of individuals with major mood disorders. Participants were recruited from a combination of a public appeal and communications campaign and via large-scale mailouts targeted at people who, based on their prescription records in the Pharmaceutical Benefits Scheme (PBS) database (i.e., having been prescribed antidepressants in the previous 4.5 years at the time). Linkage to PBS dispensing records was conducted with approval from Services Australia EREC (Reference Number MI3967), and only participants who provided explicit consent for PBS data linkage were included in analyses involving prescription records. Participants completed a comprehensive online questionnaire covering a range of health conditions, including psychological disorders and chronic pain. The study was designed to investigate the genetic and environmental factors contributing to depression, capturing detailed information on mental health, medication use, and comorbid conditions. A comprehensive description of the cohort and data collection procedures has been described previously. ^27^

### Participants

Participants were excluded if they were missing sex, age, or genotype data (n = 6,742); were of non-European ancestry (n = 6,742); reported pain for less than three months (n = 6,303); or did not use amitriptyline specifically for chronic pain (n = 5,178). Among eligible participants, those who did not answer the effectiveness question (n = 3,309) were removed from effectiveness analyses. Participants selecting “Don’t know” for effectiveness were also excluded; this exclusion may introduce selection bias if non-response is associated with metaboliser status or treatment experience. As a quality control step, we extracted Pharmaceutical Benefits Scheme (PBS) records for amitriptyline dispensation for all participants included in the analyses (n = 1,146) prior to the PharmCAT output and annotation step, to verify concordance with self-reported amitriptyline use. Following pharmacogene annotation, we additionally excluded participants who did not answer the tolerability question (n = 459) or had an indeterminate *CYP2D6* metaboliser status (n = 62) for *CYP2D6*-specific and meta-phenotype analyses. Final analytic samples included 1,146 participants for *CYP2C19*, 1,085 for *CYP2D6*, and 715 *(CYP2C19)* and 673 *(CYP2D6)* for combined meta-phenotype tolerability analyses. The full participant selection flow is shown in Figure 1.

**Figure 1.**
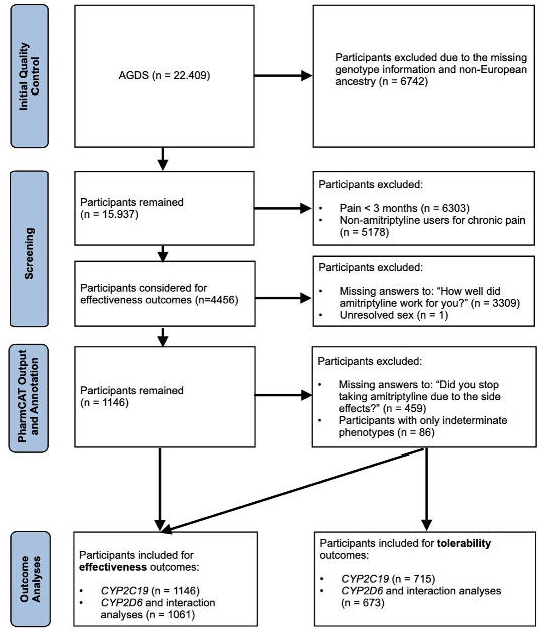
Study design and workflow. Systematic annotation of data filtering, selection, design of the study, with the number of participants.

### Genotyping and quality control

Genotyping and quality control procedures for the AGDS have been described previously. ^28^Briefly, samples were genotyped using the Illumina Global Screening Array (GSA V.2.0). Standard genotype quality control measures were applied, including exclusion of samples with >5% missingness, sex discordance, or excess heterozygosity, and removal of variants with call rate <95% or minor allele frequency <1%. Genotypes were phased and imputed using the Haplotype Reference Consortium reference panel (HRCr1.1) using the HRCr1.1 reference panel

### Pharmacogene Variant Selection

Alleles were selected based on Clinical Pharmacogenetics Implementation Consortium (CPIC) guideline recommendations for *CYP2C19*, and on the availability of the genotype array or an imputed dataset for *CYP2D6*.^29^ For *CYP2C19*, we included the clinically actionable *2 and *3 loss-of-function alleles and the *17 gain-of-function allele, which define major metaboliser phenotypes in European-ancestry populations.^18^ For *CYP2D6*, our genotyping panel captured the major loss-of-function alleles (*3, *4, *5) and the clinically relevant ultra-rapid multi-copy alleles (*1xN, *2xN), with allele definitions assigned according to PharmVar standards (Table S1).^30^

### PharmCAT output interpretation and genotype to phenotype assignment

*CYP2D6* star alleles and corresponding Gene Activity Scores (GAS) were derived from imputed dosage data using PharmCAT (Research Mode), which reports all biologically plausible diplotypes and the final GAS for each individual based on the observed variant set. Because Research Mode does not force a single metaboliser phenotype when allele ambiguity exists, PharmCAT returns all plausible diplotypes along with their associated activity scores.^31^

For participants with multiple plausible diplotype interpretations, we applied a two-step harmonisation procedure to obtain a single analytically tractable activity score: (1) Extraction of all plausible activity scores reported by PharmCAT for each participant. (2) Averaging across all plausible activity scores, thereby weighting each alternative diplotype equally and retaining the biological uncertainty inherent in the genotype call.

The final *CYP2D6* activity score was converted to a metaboliser phenotype using the CPIC genotype-to-phenotype standardisation framework. A final activity score of 0 was classified as PM, 0.25–<1.0 as IM, 1.25–2.25 as Normal metaboliser (NM), and >2.25 as UM metaboliser. ^32^ Participants with unresolved or missing activity scores after averaging were assigned an INM phenotype and excluded from downstream analyses.

*CYP2C19* matcher results and individual reports also returned multiple diplotype matches per participant.^33^ To enable quantitative analyses consistent with CYP2D6, we mapped allele-level functional categories to numeric activity values using a CPIC-compatible scheme (1.0 for normal-function alleles, 0 for no-function alleles such as *2 and *3, and 1.5 for the increased-function allele; *17. Final *CYP2C19* activity scores were generated by summing the scores by the number of alleles carried (six alleles per participant from three different SNPs tested).

Assigning numeric activity values to star alleles based on CPIC functional categories and summing them to generate an activity score is a standard practice in pharmacogenomics, as demonstrated in prior studies ^21^,^21^ and the Council of Pharmacy Standards guidelines.^34^ Metaboliser phenotypes were then assigned using prespecified activity-score cut-offs derived from the Council of Pharmacy Standards *CYP2C19* activity-score specification, as follows: AS = 0 classified as PM, 0.5 as IM, 1.0–1.5 as NM, 2.0–2.5 as RM, and ≥3.0 as UM. This approach preserved all PharmCAT-reported allelic uncertainty while ensuring consistent and standardised assignment of pharmacogene phenotypes across the cohort.

### Outcome Definitions

The primary outcomes were self-reported effectiveness and tolerability of amitriptyline in the treatment of chronic pain. Effectiveness was assessed using the question: “*How well did amitriptyline work for you?*” (Not at all well, Moderately well, Very well, Don’t know). Participants selecting ‘don’t know’ were excluded from the analysis. Tolerability was evaluated using the question: “*Did you have to stop taking amitriptyline because of side effects?*” (Yes/No).^28^ Missing or ambiguous responses were excluded.

### Statistical analyses

Associations between metaboliser phenotypes and outcomes were examined using gene-specific and combined analytical models. Effectiveness was assessed using the predefined effectiveness responses and analysed using multinomial logistic regression. Tolerability was assessed with a binary logistic regression model. For gene-specific analyses, models estimated relative risk ratios (RRR) for effectiveness outcomes and odds ratios (OR) for treatment tolerability. Models were adjusted only for age and sex. Pain severity, psychiatric comorbidities, and concomitant medications were not included as covariates because the data were collected retrospectively and were not temporally linked to amitriptyline use; medication histories were further limited by incomplete and fragmented PBS records, making accurate adjustment challenging. Including such time-ambiguous or highly collinear variables risked misclassification and overadjustment. The following reference categories were designated for the models: *not well* for effectiveness; no for tolerability; and NM type for gene-specific and gene-to-gene interactions, with concomitant NM status for both genes. Gene-stratified analyses did not assess sensitivity to the metaboliser status of the other gene. Gene-to-gene interactions were assessed for both tolerability and effectiveness outcomes by including multiplicative interaction terms in regression models. Tolerability was analysed using binary logistic regression, while effectiveness outcomes were analysed using multinomial logistic regression. For both analyses, main effects were estimated relative to normal metaboliser status for both genes. Interaction terms quantified departures from additivity of gene-specific effects; for multinomial models, one joint genotype category (*CYP2D6* PM x *CYP2C19* PM) was fixed at unity by model parameterisation and was therefore not interpreted. For all statistical analyses, both nominal and false discovery rate (FDR)-corrected p-values are reported; statistical significance was defined as pFDR < 0.05. Analyses were conducted in R (version 4.3.1) using base statistical functions and the tidyverse, data.table, and ggplot2 packages for data management and modelling. The plots were generated using Biorender (www.biorender.com).

## RESULTS

Sociodemographic, clinical, and chronic pain characteristics. The study cohort (n = 1,146) was predominantly female (79.2%), with an age of (mean [SD]): 48.8 (14.8) years, and predominantly of European descent. Among these participants, 511 had PBS-confirmed amitriptyline dispensation (codes 02417F, 02418G, 02429W), including 279 who received only one item code (mean [SD]): 2.07 (1.37) and 232 who received multiple item codes (mean [SD]): 4.57 (2.18) dispensing records, while 635 had missing data during 2013–2017. Additionally, in our questionnaire, 628 participants reported taking amitriptyline for at least one additional medical condition alongside chronic pain. Pain was assessed using a self-report questionnaire administered online via participants’ own devices, across seven main body regions, with the left and right upper and lower back (28.6%), neck and shoulders (23.6%), and hips (18.1%) most frequently affected. All participants reported one or more psychiatric comorbidities, most frequently depression (94.9%), anxiety (57.8%), and post-traumatic stress disorder (PTSD) (21.8%). The most prevalent side effects were dry mouth (33.3%), drowsiness (30.8%), and weight gain (25.8%). The average pain severity score among individuals with chronic pain was 4.7 (SE = 0.07) out of 10. The average pain intensity at the time of questionnaire being answered was 3.7 (SE 0.08). Both pain scores were calculated using a self-report questionnaire on a scale from 1 (no pain) to 10 (pain as severe as possible). There were no meaningful differences between sex stratified current and average pain intensity reporting. The full details about chronic pain characteristics, comorbidities, side effects, and average chronic pain ratings are illustrated in figures S2.A-E.

### *CYP2D6* and *CYP2C19* phenotype results

Across the cohort, *CYP2C19* and *CYP2D6* phenotypes showed substantial variability and frequent mixed or overlapping assignments. For *CYP2C19*, five phenotypes were observed (PM, IM, NM, RM, and UM), whereas *CYP2D6* showed only IM, NM, PM, and indeterminate categories. To harmonise heterogeneous phenotypic patterns, we calculated overall gene activity scores for each participant and assigned a final phenotype as described in the methods section. Following our systematic phenotype assignment, we observed three distinct phenotypes for both *CYP2C19* and *CYP2D6*: PM, IM, and NM. Those phenotype groups were used in all analyses of effectiveness and tolerability.

### Effectiveness

Descriptive analyses showed similar distributions of self-reported amitriptyline effectiveness across *CYP2D6* and *CYP2C19* metaboliser phenotypes. The proportion of participants reporting *Very well* effectiveness was similar across *CYP2D6* IM, NM, and PM (17–19%), and across *CYP2C19* phenotypes (13–20%). (Table S2.A, S2.B) *CYP2D6* metaboliser status was not meaningfully associated with reporting *moderately well* versus *not well*. Similarly, no *CYP2D6* category significantly predicted reporting *very well*. CYP2C19 IM showed a nominal trend towards lower likelihood of reporting *moderately well* versus *not well* (moderate vs poor effectiveness RRR = 0.75, 95% CI 0.55–1.02, nominal p = 0.070, pFDR = 0.280); however, this did not reach statistical significance after FDR correction. No differences were observed between PM and *very well* outcomes across any metaboliser group. (Figure 2) In interaction analyses, no statistically significant *CYP2D6–CYP2C19* effects were observed for the effectiveness outcome after FDR correction. For the moderately well outcome, point estimates were higher among combined reduced-function metaboliser groups, whereas for the very well outcome, interaction estimates tended to be lower among reduced *CYP2D6* activity combinations; however, these trends were imprecise with wide confidence intervals and are likely attributable to small subgroup sizes. (Figure 5). Given the small number of poor metabolisers in both genes (e.g., n = 24 for *CYP2C19* PM), statistical power to detect moderate interaction effects in these subgroups was limited, and the null interaction results should be interpreted as inconclusive rather than as evidence of absence (post hoc power analysis indicated detectable odds ratios ≥2.00–2.19 for *CYP2D6* poor metabolisers and ≥3.14–3.36 for *CYP2C19* poor metabolisers at 80% power).

**Figure 2.**
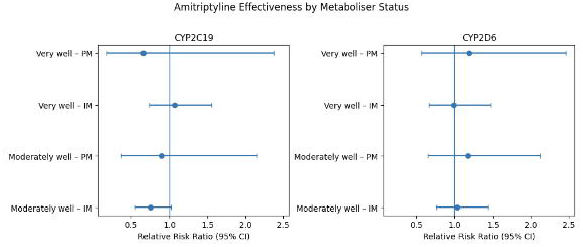
Treatment effectiveness. Forest plots show relative risk ratios (RRRs) and 95%confidence intervals for reporting *moderately well or very well* effectiveness compared with *not well*, by *CYP2C19* (left) and *CYP2D6* (right) metaboliser phenotype. Estimates were obtained from multinomial logistic regression models adjusted for age and sex, with *normal metaboliser* as the reference genotype. Although a nominal association was observed for *CYP2C19* intermediate metabolisers for the *moderately well* outcome (p = 0.070), no associations remained statistically significant after false discovery rate correction (all pFDR > 0.05). IM, intermediate metaboliser;PM, poor metaboliser.

### Tolerability

Tolerability outcomes were broadly similar across metaboliser groups, with no clear differences in discontinuation due to side effects across *CYP2D6* or *CYP2C19* phenotypes. The distribution of responses across metaboliser phenotypes is reported in detail in tables S2.C and S2.D. Logistic regression showed no significant statistical associations between *CYP2D6* metaboliser status and stopping amitriptyline due to side effects. *CYP2C19* IM showed nominally lower odds of stopping amitriptyline due to side effects compared with NM (OR 0.66, 95% CI 0.48–0.92, nominal p = 0.013, pFDR = 0.064), though this association did not survive correction for multiple testing. PM showed no significant difference (Figure 3).

**Figure 3.**
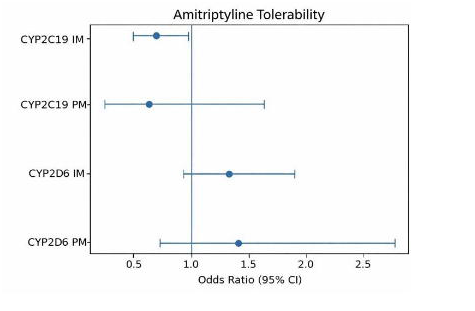
Treatment tolerability. Forest plot of odds ratios (ORS) and 95% confidence intervals fordiscontinuation of amitriptyline due to side effects by *CYP2C19* and *CYP2D6* metaboliser status. Models were adjusted for age and sex, with *normal metaboliser* as the reference genotype. Aftercorrection for multiple testing, no associations reached statistical significance (*CYP2C19* intermediate metaboliser: pFDR = 0.127; *CYP2C19* poor metaboliser: pFDR = 0.422; *CYP2D6* intermediate metaboliser: pFDR = 0.244; *CYP2D6* poor metaboliser: pFDR = 0.422). IM,intermediate metaboliser; PM, poor metaboliser; NM, normal metaboliser.

Interaction analyses showed that no *CYP2D6* or *CYP2C19* metaboliser phenotype was significantly associated with discontinuation due to side effects after FDR. *CYP2C19* IM showed a nominal trend towards reduced odds of discontinuation, whereas *CYP2D6* reduced-function metabolisers showed point estimates suggesting increased odds; however, these trends were imprecise and did not reach statistical significance. (Figure 4)

**Figure 4.**
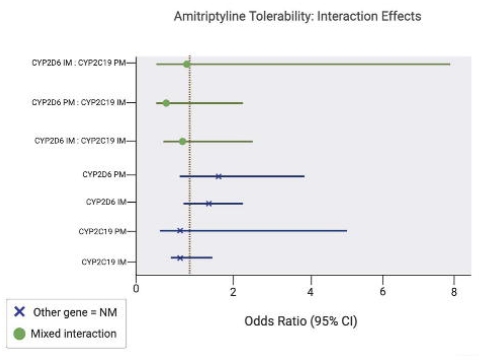
Treatment tolerability (interactions). Associations between *CYP2D6* and *CYP2C19* metaboliser phenotypes and amitriptyline discontinuation due to side effects. Main effects (whenthe other gene is NM): *CYP2C19* IM (OR 0.74, 95% CI 0.37–1.47; PFDR=0.65), *CYP2C19* PM (0.75, 0.09–4.80; PFDR–0.76), *CYP2D6* IM (1.33, 0.86–2.05; pFDR–0.36), and *CYP2D6* PM(1.77, 0.78–4.15; PFDR–0.36). Interaction terms: *CYP2D6* IM×*CYP2C19* IM (0.98, 0.44–2.20;PFDR–0.96), *CYP2D6* PM *CYP2C19* IM (0.45, 0.10–2.01; pFDR–0.30), and *CYP2D6* IM×*CYP2C79* PM (0.90, 0.11–9.04; pFDR–0.92).

**Figure 5.**
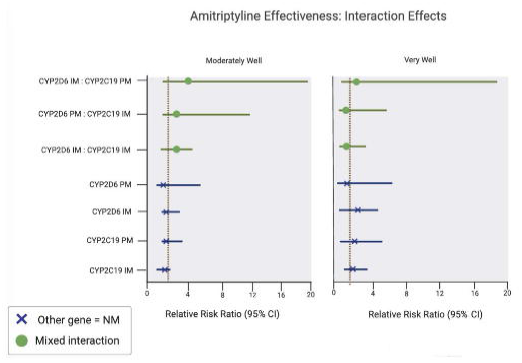
Treatment effectiveness (interactions). Associations between *CYP2D6* and *CYP2C19* metaboliser phenotypes and amitriptyline effectiveness outcomes. Moderately well: *CYP2D6* IM(RRR 1.05, 95% CI 0.73–1.51; pFDR–1), *CYP2D6* PM (0.99, 0.49–1.98; pFDR=1), *CYP2C19* IM(0.78, 0.41–1.48; pFDR–0.814), *CYP2C19* PM (0.47, 0.09–2.50; pFDR–0.814); interactions:*CYP2D6* IM×*CYP2C19* IM (0.90, 0.43–1.88; pFDR=1), *CYP2D6* PM×*CYP2C19* IM (1.78, 0.47–6.73; PFDR–0.814), and *CYP2D6* IM×*CYP2C19* PM (2.55, 0.35–18.60; PFDR–0.814). Very well:*CYP2D6* IM (1.16, 0.71–1.89; pFDR–0.693), *CYP2D6* PM (1.49, 0.65–3.46; pFDR–0.497),*CYP2C19* IM (1.63, 0.79–3.39; pFDR–0.497), *CYP2C19* PM (0.58, 0.06–5.24; pFDR=0.700);interactions: *CYP2D6* IM×*CYP2C19* IM (0.58, 0.25–1.38; pFDR–0.497), *CYP2D6* PM×*CYP2C19* IM (0.36, 0.06–2.34; pFDR–0.497), and *CYP2D6* IM×*CYP2C19* PM (1.29, 0.09–19.57;pFDR–0.853).

## DISCUSSION

In this large cohort of amitriptyline users from the AGDS study, neither *CYP2C19* nor *CYP2D6* metaboliser status showed strong or consistent associations with self-reported treatment effectiveness or tolerability. These essentially null associations are broadly consistent with the current consensus in pharmacogenomics: although *CYP2C19* and *CYP2D6* variation clearly influences TCA pharmacokinetics, its impact on clinical outcomes tends to be modest at the population level. We observed no strong relationship between metaboliser phenotype and patient-reported benefit, and only nominal associations for *CYP2C19* IM (modestly lower odds of discontinuation and of reporting moderate effectiveness), neither of which survived FDR correction. We interpret these findings as reflecting the attenuation of pharmacogenomic effects in a real-world chronic pain context characterised by low-dose prescribing, clinical titration, polypharmacy, and heterogeneous pain mechanisms.

Our findings differ from small therapeutic drug-monitoring studies, in which *CYP2C19/CYP2D6* genotypes have been used to identify individuals at elevated risk of adverse reactions during amitriptyline therapy ^22,35^, and from a recent cross-generational electronic health record analysis reporting enrichment of *CYP2C19* PM among families with repeated early amitriptyline discontinuation.^36^ However, those studies focused on extreme metaboliser groups, relied on selected clinical populations, or examined antidepressant treatment more broadly, rather than limiting their findings to chronic pain subjects, as we demonstrated in our study. Our cohort consisted mainly of intermediate and normal metabolisers with chronic pain and high psychiatric comorbidity, particularly depression and anxiety. In this real-world setting, characterised by polypharmacy, lower TCA doses for pain, and uncontrolled heterogeneous pain mechanisms, genotype–phenotype associations may be attenuated and more difficult to detect. The retrospective design of AGDS and the absence of detailed dosing information may have further diluted observable signals. Adittionally more than half of the analysis sample were prescribed amitriptyline for more than one medical condition, which limits our ability to consider this cohort as exclusively prescribed amitriptyline for chronic pain.

### Dose–effect threshold and low-dose prescribing

A critical consideration for interpreting our null findings is the dose range at which amitriptyline is used for chronic pain. The relatively small number of dispensing events per participant across three amitriptyline dosages within the 5-year time window, therefore, suggests intermittent or low-dose use in individuals with valid amitriptyline dispensing data (n=511), consistent with real-world chronic pain prescribing practices. Pharmacokinetic studies demonstrating clinically relevant CYP-mediated differences in amitriptyline and nortriptyline plasma concentrations have typically employed antidepressant-range doses (75–150 mg/day), at which metabolic variation can produce supra- or sub-therapeutic plasma levels ^37^,^22^. In contrast, chronic pain prescribing in Australia and comparable settings typically involves doses of 10–50 mg/day ^38^, often remaining well below the concentration threshold at which metabolic differences between normal and poor metabolisers would be expected to trigger toxicity or therapeutic failure. Under these conditions, even a poor metaboliser may not accumulate sufficient drug to experience clinically distinguishable adverse effects beyond what a normal metaboliser would experience. This dose–effect consideration is arguably the most parsimonious explanation for why our pharmacogenomic associations were attenuated, and underscores the importance of accounting for dose context when extrapolating pharmacokinetic evidence to clinical outcomes.

### Mechanistic considerations: nociplastic pain and metabolite profiles

Amitriptyline’s analgesic mechanism in chronic pain is thought to involve descending inhibitory pathway modulation via serotonin and noradrenaline reuptake inhibition, which is mechanistically distinct from its antidepressant action. The relative contributions of amitriptyline (a more potent serotonin reuptake inhibitor) and its demethylated metabolite nortriptyline (a more potent noradrenaline reuptake inhibitor) to pain relief are not fully understood. *CYP2C19* preferentially catalyses the N-demethylation of amitriptyline to nortriptyline, and variation in *CYP2C19* activity could therefore shift the amitriptyline-to-nortriptyline ratio, potentially influencing which neurotransmitter pathway predominates. Whether such shifts are clinically meaningful at the low doses used for pain remains uncertain, but this pharmacological complexity may contribute to the heterogeneous and largely null associations observed in our study. In particular, for patients with nociplastic pain—where central sensitisation rather than peripheral nociceptive input drives symptoms—the relevant pharmacodynamic targets may be less sensitive to the metabolic variation captured by CYP genotyping ^39^.

### Titration-based prescribing as implicit pharmacogenomic management

Contemporary prescribing guidelines for amitriptyline in chronic pain recommend initiating at low doses with gradual upward titration guided by clinical response and tolerability. This approach effectively functions as empirical pharmacogenomic management: patients who are slow metabolisers tend to experience side effects earlier in the titration process and are maintained at lower doses or switched to alternatives, while rapid metabolisers may be titrated to higher doses without encountering toxicity. In a retrospective cohort such as ours, where dosing trajectories are unobserved, this clinical “self-correction” may substantially attenuate the relationship between genotype and outcome, because the prescribing process has already compensated for metabolic variation. This interpretation is consistent with emerging evidence suggesting that pharmacogenomic testing may offer the greatest incremental value when standard titration is impractical, when higher doses are required, or when prior treatment failure suggests an extreme metaboliser phenotype ^35^.

Although amitriptyline metabolism is strongly influenced by *CYP2C19* and *CYP2D6* activity in in vitro and pharmacokinetic studies ^40,41^, translating these findings to clinical outcomes remains complex. The chronic pain response to amitriptyline is shaped by multiple interacting factors, including chronic pain subtypes, psychological comorbidity, placebo and expectation effects, dose titration, adherence, and environmental influences, any of which may overshadow genetic contributions. ^42^

Nonetheless, pharmacogenetic testing remains clinically useful for identifying individuals at the extremes of *CYP2C19* or *CYP2D6* function, where the risk of substantial over- or under-exposure to amitriptyline is greatest. However, its incremental benefit for routine prescribing in primary care, where most chronic pain is managed, may be limited.^43^ In these settings, patients are typically initiated on low doses with gradual titration, allowing side effects or lack of efficacy to be identified through standard clinical monitoring. Under such conditions, careful dose adjustment, patient education, and regular follow-up may offer greater practical benefit than broad pharmacogenetic screening. Genotyping may be most informative when higher TCA doses are considered or when prior intolerance suggests an extreme metaboliser phenotype for chronic pain.

Importantly, our findings do not fully contradict existing guideline recommendations. Rather, they refine their interpretation. Current CPIC and international guidelines emphasise genotype-informed avoidance or dose adjustment for extreme metaboliser phenotypes (PM or UM), but acknowledge weaker evidence for predicting treatment effectiveness or tolerability among IM and NM. ^29, 44, 45^ Given that PMs were rare in our sample, our results support the view that genotype-informed dose adjustments may have only modest effects on average population-level outcomes. A key limitation reinforcing this interpretation is the inability to reconstruct individual dosing trajectories from PBS records. This reflects incomplete linkage consent or prescription coverage within the 2013–2017 observation window, as well as inherent characteristics of PBS dispensing data which is not designed to capture precise daily dosing, titration schedules, and adherence.

A strength of this study is its rigorous and scalable methodological framework. We implemented automated allele-to-phenotype translation using PharmCAT in research mode for *CYP2D6* and integrated these outputs with a structured statistical decision algorithm to derive final metaboliser assignments. Although PharmCAT research mode is not yet available for *CYP2C19*, its future implementation would further enhance genotype-to-phenotype harmonisation, particularly given the prevalence of structural variants in this gene.^46^ Our hybrid approach reduces the risk of manual misclassification, improves reproducibility across large genomic datasets, and appropriately accounts for uncertainty arising from CNVs and SVs, an especially important consideration for *CYP2D6*.^47^ In addition, our gene-to-gene interaction analyses offer a novel framework for evaluating the combined pharmacogenetic effects of *CYP2C19* and *CYP2D6*, which jointly contribute to amitriptyline metabolism, rather than examining each gene in isolation. To our knowledge, this is also the largest study to date to investigate amitriptyline pharmacogenetics in a cohort with co-occurring chronic pain and depression.

Overall, our findings underscore the need for nuanced expectations regarding the predictive value of pharmacogenomics for amitriptyline response in chronic pain. While *CYP2C19* and *CYP2D6* genotyping remains clinically valuable for identifying individuals at risk of extreme drug exposure, its utility for predicting self-reported effectiveness or tolerability in unselected, community-based chronic pain populations appears limited. Future research should prioritise prospective designs with documented dosing trajectories, measurement of plasma drug concentrations, and assessment of pharmacodynamic biomarkers alongside refined chronic pain phenotyping (including nociplastic subtypes) and longitudinal outcome measures. Such studies may better delineate the biological and clinical pathways through which genetic variation influences treatment response, and clarify the settings in which pharmacogenomic testing offers meaningful clinical value beyond standard titration-based prescribing.

## Data Availability

All data produced and used in the present study are available upon reasonable request to the authors.

## ETHICS STATEMENT

The Australian Genetics of Depression Study (AGDS) was approved by the QIMR Berghofer Medical Research Institute Research Ethics Committee (P2118) and by Services Australia External Request Evaluation Committee (EREC), which govern data linkage and access. All participants provided written informed consent for participation, including consent for genetic analyses and linkage to administrative health records where applicable.

## AUTHOR’S CONTRIBUTIONS

NGM led overall study design and data collection for AGDS; GEF and MER conceived this study and jointly supervised its conduct; BU performed the statistical analyses and led drafting of the first version of the manuscript with support from NSO and SDT; SEM and PAL obtained, processed and analysed the PBS data; all authors were involved in the drafting of the manuscript, provided feedback, and approved the final version before submission.

## ACKNOWLEDGMENTS

BU is supported by the QUT TFS International Tuition Fee Scholarship and a QIMR Computational Neurogenomics Scholarship. MER gratefully acknowledges fellowship support from the Rebecca L. Cooper Medical Research Foundation (F20231230), awarded through the Al & Val Rosenstruss Fellowship. The AGDS was primarily funded by Australia’s National Health and Medical Research Council (NHMRC), grant (APP1086683).GF is funded by an NHMRC Emerging Leadership Investigator Fellowship (APP2009808). SEM is funded by an NHMRC Leadership grant (APP2025674).

## DECLARATION OF GENERATIVE ARTIFICIAL INTELLIGENCE (AI) IN SCIENTIFIC WRITING

We declare no involvement of AI in the data analysis, preparation, and editing of this manuscript.

## REFERENCES

1. Curatolo M. Central sensitization and pain: Pathophysiologic and clinical insights. Curr Neuropharmacol [Internet] Bentham Science Publishers Ltd.; 2024; 22: 15–22 Available from: 10.2174/1570159X20666221012112725

2. Nekovarova T, Yamamotova A, Vales K, Stuchlik A, Fricova J, Rokyta R. Common mechanisms of pain and depression: are antidepressants also analgesics? Front Behav Neurosci [Internet] Frontiers Media SA; 2014; 8: 99 Available from: 10.3389/fnbeh.2014.00099

3. Kang Y, Trewern L, Jackman J, McCartney D, Soni A. Chronic pain: definitions and diagnosis. BMJ [Internet] British Medical Journal Publishing Group; 2023; 381: e076036 Available from: 10.1136/bmj-2023-076036

4. Chronic pain in Australia [Internet]. Australian Institute of Health and Welfare. [cited 2025 Oct 19]. Available from: https://www.aihw.gov.au/reports/chronic-disease/chronic-pain-in-australia

5. Wong J, Motulsky A, Eguale T, Buckeridge DL, Abrahamowicz M, Tamblyn R. Treatment indications for antidepressants prescribed in primary care in Quebec, Canada, 2006-2015. JAMA [Internet] American Medical Association (AMA); 2016; 315: 2230 Available from: 10.1001/jama.2016.3445

6. Tamblyn R, Bates DW, Buckeridge DL, et al. Multinational comparison of new antidepressant use in older adults: a cohort study. BMJ Open [Internet] BMJ; 2019; 9: e027663 Available from: 10.1136/bmjopen-2018-027663

7. Coetzee R, Johnson Y, van Niekerk J, Namane M. Amitriptyline prescribing in public sector healthcare facilities in the Western Cape, South Africa. PLoS One [Internet] Public Library of Science (PLoS); 2020; 15: e0231675 Available from: 10.1371/journal.pone.0231675

8. Gisev N, Nielsen S, Campbell G, et al. Antidepressant use among people prescribed opioids for chronic noncancer pain. Pain Med [Internet] Oxford University Press (OUP); 2019; 20: 2450–8 Available from: 10.1093/pm/pnz009

9. Ferreira GE, Di Donato M, Maher CG, Shaheed CA, Mathieson S, Collie A. Patterns of antidepressant use in people with low back pain: A retrospective study using workers’ compensation data. Eur J Pain [Internet] Wiley; 2025; 29: e4773 Available from: 10.1002/ejp.4773

10. van den Driest JJ, Schiphof D, de Wilde M, Bindels PJE, van der Lei J, Bierma-Zeinstra SMA. Antidepressant and anticonvulsant prescription rates in patients with osteoarthritis: a population-based cohort study. Rheumatology (Oxford) [Internet] Oxford University Press (OUP); 2021; 60: 2206–16 Available from: 10.1093/rheumatology/keaa544

11. Riediger C, Schuster T, Barlinn K, Maier S, Weitz J, Siepmann T. Adverse effects of antidepressants for chronic pain: A systematic review and meta-analysis. Front Neurol [Internet] Frontiers Media SA; 2017; 8: 307 Available from: 10.3389/fneur.2017.00307

12. Hudson B, Williman JA, Stamp LK, et al. Nortriptyline for pain in knee osteoarthritis: a double-blind randomised controlled trial in New Zealand general practice. Br J Gen Pract [Internet] Royal College of General Practitioners; 2021; 71: e538–46 Available from: 10.3399/BJGP.2020.0797

13. Moore RA, Derry S, Aldington D, Cole P, Wiffen PJ. Amitriptyline for neuropathic pain in adults. Cochrane Database Syst Rev [Internet] Wiley; 2015; 2019: CD008242 Available from: 10.1002/14651858.CD008242.pub3

14. Moore RA, Derry S, Aldington D, Cole P, Wiffen PJ. Amitriptyline for neuropathic pain and fibromyalgia in adults. Cochrane Database Syst Rev [Internet] 2012; 12: CD008242 Available from: 10.1002/14651858.CD008242.pub2

15. Brueckle M-S, Thomas ET, Seide SE, et al. Amitriptyline’s anticholinergic adverse drug reactions-A systematic multiple-indication review and meta-analysis. PLoS One [Internet] PLoS One; 2023; 18: e0284168 Available from: 10.1371/journal.pone.0284168

16. Ryu S, Park S, Lee JH, et al. A study on CYP2C19 and CYP2D6 polymorphic effects on pharmacokinetics and pharmacodynamics of amitriptyline in healthy Koreans. Clin Transl Sci [Internet] 2017; 10: 93–101 Available from: 10.1111/cts.12451

17. Grasmäder K, Verwohlt PL, Rietschel M, et al. Impact of polymorphisms of cytochrome-P450 isoenzymes 2C9, 2C19 and 2D6 on plasma concentrations and clinical effects of antidepressants in a naturalistic clinical setting. Eur J Clin Pharmacol [Internet] Springer Science and Business Media LLC; 2004; 60: 329–36 Available from: 10.1007/s00228-004-0766-8

18. Shubbar Q, Alchakee A, Issa KW, Adi AJ, Shorbagi AI, Saber-Ayad M. From genes to drugs: CYP2C19 and pharmacogenetics in clinical practice. Front Pharmacol [Internet] Frontiers Media SA; 2024; 15: 1326776 Available from: 10.3389/fphar.2024.1326776

19. Mifsud Buhagiar L, Casha M, Grech A, Serracino Inglott A, LaFerla G. The interplay between pharmacogenetics, concomitant drugs and blood levels of amitriptyline and its main metabolites. Per Med [Internet] Informa UK Limited; 2022; 19: 113–23 Available from: 10.2217/pme-2021-0022

20. Brouwer JMJL, Wardenaar KJ, Nolte IM, et al. Association of CYP2D6 and CYP2C19 metabolizer status with switching and discontinuing antidepressant drugs: an exploratory study. BMC Psychiatry [Internet] Springer Science and Business Media LLC; 2024; 24: 394 Available from: 10.1186/s12888-024-05764-6

21. Li D, Pain O, Fabbri C, et al. Metabolic activity of CYP2C19 and CYP2D6 on antidepressant response from 13 clinical studies using genotype imputation: a meta-analysis. Transl Psychiatry [Internet] Springer Science and Business Media LLC; 2024; 14: 296 Available from: 10.1038/s41398-024-02981-1

22. Steimer W, Zöpf K, von Amelunxen S, et al. Amitriptyline or not, that is the question: pharmacogenetic testing of CYP2D6 and CYP2C19 identifies patients with low or high risk for side effects in amitriptyline therapy. Clin Chem [Internet] Oxford University Press (OUP); 2005; 51: 376–85 Available from: 10.1373/clinchem.2004.041327

23. Chaudhry M, Alessandrini M, Rademan J, et al. Impact of CYP2D6 genotype on amitriptyline efficacy for the treatment of diabetic peripheral neuropathy: a pilot study. Pharmacogenomics [Internet] Informa UK Limited; 2017; 18: 433–43 Available from: 10.2217/pgs-2016-0185

24. Sylwander C, Larsson I, Andersson M, Bergman S. The impact of chronic widespread pain on health status and long-term health predictors: a general population cohort study. BMC Musculoskelet Disord [Internet] Springer Science and Business Media LLC; 2020; 21: 36 Available from: 10.1186/s12891-020-3039-5

25. Fayaz A, Croft P, Langford RM, Donaldson LJ, Jones GT. Prevalence of chronic pain in the UK: a systematic review and meta-analysis of population studies. BMJ Open [Internet] BMJ Open; 2016; 6: e010364 Available from: 10.1136/bmjopen-2015-010364

26. Kosek E. The concept of nociplastic pain-where to from here? Pain [Internet] Ovid Technologies (Wolters Kluwer Health); 2024; 165: S50–7 Available from: 10.1097/j.pain.0000000000003305

27. Byrne EM, Kirk KM, Medland SE, et al. Cohort profile: the Australian genetics of depression study. BMJ Open [Internet] BMJ; 2020; 10: e032580 Available from: 10.1136/bmjopen-2019-032580

28. Campos AI, Byrne EM, Mitchell BL, et al. Impact of CYP2C19 metaboliser status on SSRI response: a retrospective study of 9500 participants of the Australian Genetics of Depression Study. Pharmacogenomics J [Internet] Springer Science and Business Media LLC; 2022; 22: 130–5 Available from: 10.1038/s41397-022-00267-7

29. Hicks JK, Sangkuhl K, Swen JJ, et al. Clinical pharmacogenetics implementation consortium guideline (CPIC) for CYP2D6 and CYP2C19 genotypes and dosing of tricyclic antidepressants: 2016 update. Clin Pharmacol Ther [Internet] 2017; 102: 37–44 Available from: 10.1002/cpt.597

30. Pratt VM, Cavallari LH, Del Tredici AL, et al. Recommendations for clinical CYP2D6 genotyping allele selection: A joint consensus recommendation of the Association for molecular pathology, college of American pathologists, dutch pharmacogenetics Working Group of the royal dutch pharmacists Association, and the European society for pharmacogenomics and personalized therapy. J Mol Diagn [Internet] Elsevier BV; 2021; 23: 1047–64 Available from: 10.1016/j.jmoldx.2021.05.013

31. Sangkuhl K, Whirl-Carrillo M, Whaley RM, et al. Pharmacogenomics Clinical Annotation Tool (PharmCAT). Clin Pharmacol Ther [Internet] Wiley; 2020; 107: 203–10 Available from: 10.1002/cpt.1568

32. Caudle KE, Sangkuhl K, Whirl-Carrillo M, et al. Standardizing CYP2D6 genotype to phenotype translation: Consensus recommendations from the Clinical Pharmacogenetics Implementation Consortium and Dutch Pharmacogenetics Working Group. Clin Transl Sci [Internet] Wiley; 2020; 13: 116–24 Available from: 10.1111/cts.12692

33. Tibben BM, Gaedigk A, Gong L, et al. The Clinical Pharmacogenetics Implementation Consortium’s consensus-based framework for assigning allele function. Am J Hum Genet [Internet] Elsevier BV; 2025; Available from: 10.1016/j.ajhg.2025.10.004

34. 5.2 Activity Scores and Phenotype Prediction – Council on Pharmacy Standards [Internet]. [cited 2026 Feb 17]. Available from: https://pharmacystandards.org/cpxp/section-5-2-activity-scores-and-phenotype-prediction/?utm_source

35. Steimer W, Zöpf K, von Amelunxen S, et al. Allele-specific change of concentration and functional gene dose for the prediction of steady-state serum concentrations of amitriptyline and nortriptyline in CYP2C19 and CYP2D6 extensive and intermediate metabolizers. Clin Chem [Internet] Oxford University Press (OUP); 2004; 50: 1623–33 Available from: 10.1373/clinchem.2003.030825

36. Magavern EF, Marengo G, Megase M, Smedley D, Caulfield MJ. Family health history and pharmacogenomics show cross generation premature amitriptyline discontinuation is associated with CYP2C19 loss of-function enrichment. Commun Med (Lond) [Internet] Springer Science and Business Media LLC; 2025; 5: 437 Available from: 10.1038/s43856-025-01156-3

37. Baumann P, Jonzier-Perey M, Koeb L, Lê PK, Tinguely D, Schöpf J. Amitriptyline pharmacokinetics and clinical response: I. Free and total plasma amitriptyline and nortriptyline. Int Clin Psychopharmacol [Internet] Int Clin Psychopharmacol; 1986; 1: 89–101 Available from: 10.1097/00004850-198604000-00001

38. Amitriptyline for nerve pain [Internet]. NPS MedicineWise. 2018 [cited 2026 Feb 16]. Available from: https://www.nps.org.au/consumers/amitriptyline-for-nerve-pain

39. Obata H. Analgesic mechanisms of antidepressants for neuropathic pain. Int J Mol Sci [Internet] Multidisciplinary Digital Publishing Institute; 2017; 18: 2483 Available from: 10.3390/ijms18112483

40. Jornil J, Linnet K. Roles of polymorphic enzymes CYP2D6 and CYP2C19 for in vitro metabolism of amitriptyline at therapeutic and toxic levels. Forensic Toxicol [Internet] Springer Science and Business Media LLC; 2009; 27: 12–20 Available from: 10.1007/s11419-008-0063-9

41. Koh A, Pak KC, Choi HY, et al. Quantitative modeling analysis demonstrates the impact of CYP2C19 and CYP2D6 genetic polymorphisms on the pharmacokinetics of amitriptyline and its metabolite, nortriptyline. J Clin Pharmacol [Internet] SAGE Publications; 2019; 59: 532–40 Available from: 10.1002/jcph.1344

42. Su M. Amitriptyline Therapy in Chronic Pain. Int Arch Clin Pharmacol [Internet] ClinMed International Library; 2015; 1 Available from: 10.23937/2572-3987.1510001

43. Smith DM, Beyene R, Kolm P, et al. A randomized hybrid-effectiveness trial comparing pharmacogenomics (PGx) to standard care: The PGx applied to chronic pain treatment in primary care (PGx-ACT) trial. Clin Transl Sci [Internet] Wiley; 2025; 18: e70154 Available from: 10.1111/cts.70154

44. Vos CF, Ter Hark SE, Schellekens AFA, et al. Effectiveness of genotype-specific tricyclic antidepressant dosing in patients with major depressive disorder: A randomized clinical trial. JAMA Netw Open [Internet] JAMA Netw Open; 2023; 6: e2312443 Available from: 10.1001/jamanetworkopen.2023.12443

45. Hicks JK, Swen JJ, Thorn CF, et al. Clinical Pharmacogenetics Implementation Consortium guideline for CYP2D6 and CYP2C19 genotypes and dosing of tricyclic antidepressants. Clin Pharmacol Ther [Internet] Wiley; 2013; 93: 402–8 Available from: https://files.cpicpgx.org/data/guideline/publication/TCA/2013/23486447.pdf

46. Scott SA, Martis S, Peter I, Kasai Y, Kornreich R, Desnick RJ. Identification of CYP2C19*4B: pharmacogenetic implications for drug metabolism including clopidogrel responsiveness. Pharmacogenomics J [Internet] Springer Science and Business Media LLC; 2012; 12: 297–305 Available from: 10.1038/tpj.2011.5

47. Lopes JL, Harris K, Karow MB, et al. Targeted genotyping in clinical pharmacogenomics: What is missing? J Mol Diagn [Internet] Elsevier BV; 2022; 24: 253–61 Available from: 10.1016/j.jmoldx.2021.11.008

