## Supplementary Material for "Evaluating the Clinical Impact of CYP2C19 and CYP2D6 on Amitriptyline Outcomes in a Real-World Chronic Pain Cohort"

**Supplementary Appendix 1**

Table of Contents

Supplementary Tables………………………………..2

Supplementary Table 1……………………………….2

Supplementary Table 2……………………………….3

Supplementary Table 3A and 3B……………………..3

Supplementary Table 4A and 4B……………………..4

Supplementary Figures……………………………….5

Supplementary Figure 1A and 1B……………………5

Supplementary Figure 2A and 2B……………………6

Supplementary Figure 3………...……………………7

**Supplementary Table 1.** European allele-frequency distribution of CYP2D6 star alleles (A). Frequencies are obtained from the [CPIC/PharmVar allele-frequency panel](https://www.clinpgx.org/page/cyp2d6RefMaterials), which compiles weighted averages from CYP2D6-specific genotyping, sequencing, and copy-number assays reported across multiple European ancestry cohorts.

| **rsID** | **Star Allele(s)** | **Function** | **EUR Frequency** |
| --- | --- | --- | --- |
| rs3892097 | *4 | No function | 0.18485 |
| rs16947 | *2, *35, *41 | Normal / decreased | 0.18536; 0.05468; 0.09238 |
| rs1135840 | *2, *17, *29, *35, *41 | Normal / decreased | 0.18536; 0.00392; 0.00105; 0.05468; 0.09238 |
| rs1058172 | *2, *35, *41 | Normal / decreased | 0.18536; 0.05468; 0.09238 |
| rs1065852 | *10, *36 | Decreased | 0.01571; 0.000175 |
| rs1135822 | *1 / *2 backbone | Normal | 0.2850; 0.18536 |
| rs1135828 | *1 / *2 backbone | Normal | 0.2850; 0.18536 |
| rs5030865 | *4 suballele | No function | 0.18485 |
| rs5030867 | *4 suballele | No function | 0.18485 |
| rs28371696 | *36 | No function | 0.000175 |
| rs118203758; rs138100349; rs150163869; rs28371706; rs28371710; rs28371717; rs28371725; rs141009491; rs267608319; rs28371733; rs369177208; rs373813287; rs535642512; rs569439709; rs59421388; rs61736512; rs72549358; rs75386357; rs769258; rs77913725; rs78209835; rs78482768; rs79292917 | Rare CYP2D6 alleles (*117–*163 or unassigned) | Not known | Each < 0.001 |

**Supplementary Table 2.** Carrier frequency of *CYP2D6* star alleles in the total cohort (n=1146) diplotype solution space. Frequencies represent the proportion of participants with at least one candidate diplotype containing each allele.

| **Functional class** | **Star allele** | **Participants (N)** | **Carrier frequency (%)** |
| --- | --- | --- | --- |
| **Normal function** | *2 | 815 | 71.05 |
| **Normal function** | *1 | 375 | 32.69 |
| **No-function** | *4 | 359 | 31.30 |
| **Rare alleles (no function or unassigned)** | *117–*163 (grouped) | 964 | 84.05 |

**Supplementary Table 3.** (A) Patient-reported treatment effectiveness by CYP2D6 metaboliser phenotype and (B) by CYP2C19 metaboliser phenotype. Effectiveness was categorised as “Not well”, “Moderately well”, or “Very well”. Percentages represent the proportion of the total analysed cohort within each gene-specific analysis, rather than row-wise percentages within metaboliser phenotype groups.

**A)**

| **Metaboliser phenotype (CYP2D6)** | **N total per phenotype** | **Not well** | **Moderately well** | **Very well** |
| --- | --- | --- | --- | --- |
| **Intermediate** | 752 | 350 (46.5%) | 272 (36.2%) | 130 (17.3%) |
| **Normal** | 263 | 125 (47.5%) | 92 (35.0%) | 46 (17.5%) |
| **Poor** | 70 | 31 (44.3%) | 26 (37.1%) | 13 (18.6%) |

**B)**

| **Metaboliser phenotype (CYP2C19)** | **N total per phenotype** | **Not well** | **Moderately well** | **Very well** |
| --- | --- | --- | --- | --- |
| **Poor** | 24 | 12 (50.0%) | 9 (37.5%) | 3 (12.5%) |
| **Intermediate** | 297 | 146 (49.2%) | 92 (31.0%) | 59 (19.9%) |
| **Normal** | 826 | 373 (45.2%) | 316 (38.3%) | 137 (16.6%) |

**Supplementary Table 4.** (A) Treatment tolerability by CYP2D6 metaboliser phenotype and (B) by CYP2C19 metaboliser phenotype, defined as continuation (“Did not stop”) or discontinuation due to side-effects (“Stopped”). Percentages represent the proportion of the total analysed cohort within each gene-specific analysis, rather than row-wise percentages within metaboliser phenotype groups.

**A)**

| **Metaboliser phenotype (CYP2D6)** | **N total per phenotype** | **Did not stop** | **Stopped (side-effects)** |
| --- | --- | --- | --- |
| **Intermediate** | 467 | 212 (28.2%) | 255 (33.9%) |
| **Normal** | 162 | 85 (32.3%) | 77 (29.3%) |
| **Poor** | 44 | 19 (27.1%) | 25 (35.7%) |

**B)**

| **Metaboliser phenotype (CYP2C19)** | **N total per pehnotype** | **Did not stop** | **Stopped (side-effects)** |
| --- | --- | --- | --- |
| **Poor** | 18 | 10 (41.7%) | 8 (33.3%) |
| **Intermediate** | 200 | 108 (36.4%) | 92 (31.0%) |
| **Normal** | 497 | 218 (26.4%) | 279 (33.8%) |

**Supplementary Figure 1.** Colour-coded illustration of reported overlapping chronic pain sites (A), percentage frequencies of chronic pain across body sites, and the reported pain sites are overlapping across the cohort (B**)**.

**A)**

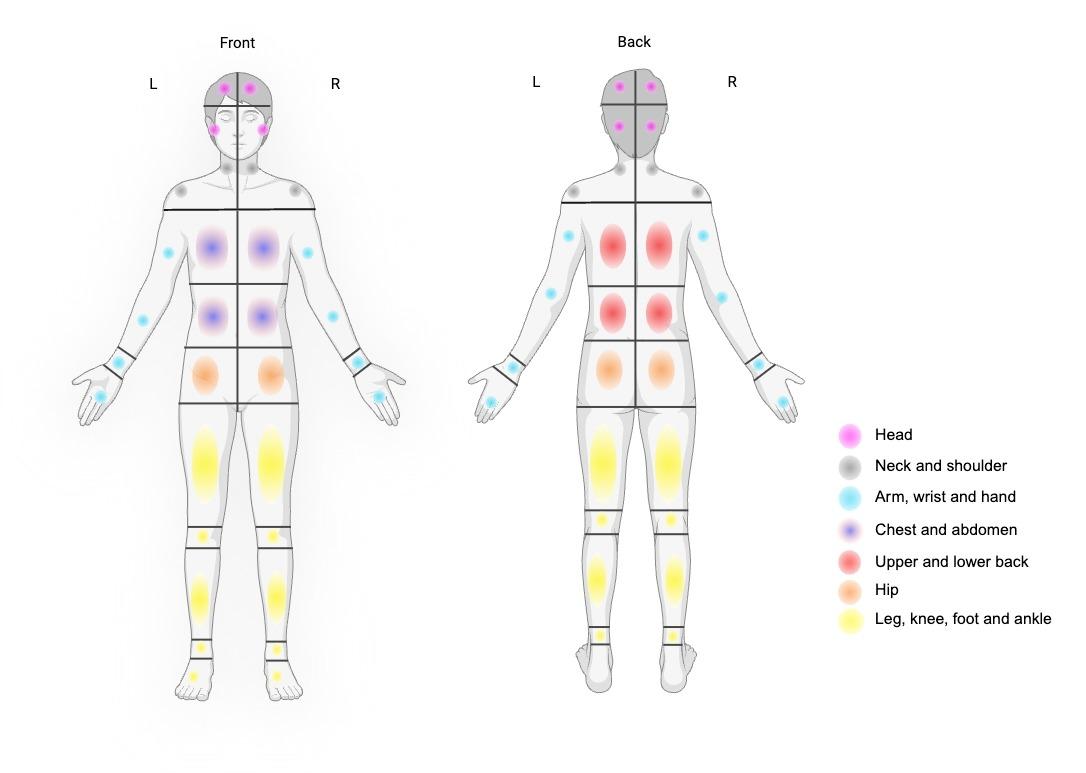

**B)**

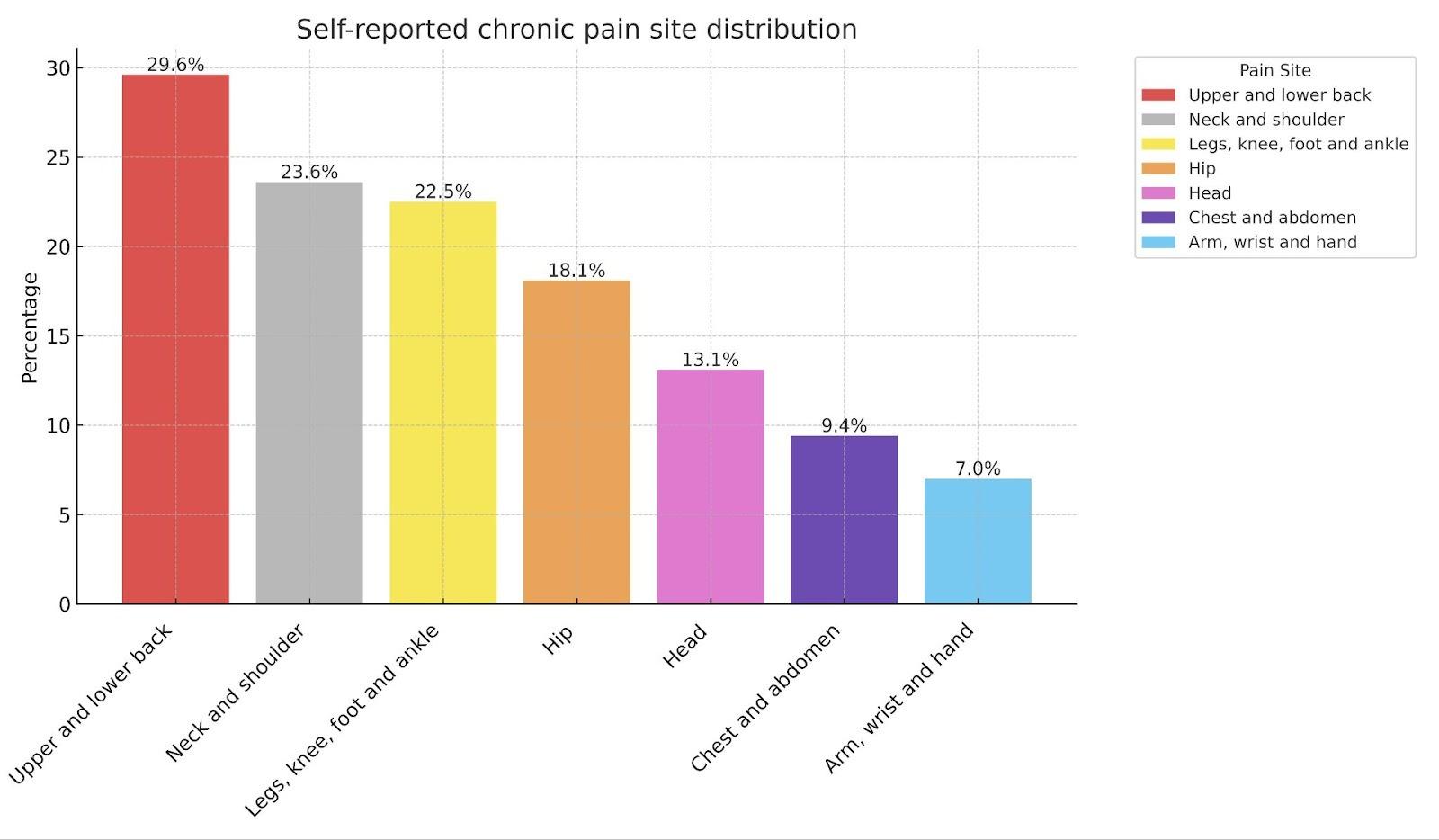

**Supplementary Figure 2.** Overall, participants with diagnosed psychiatric comorbidities reported at least one psychiatric condition (A). Side effects reported in participants taking amitriptyline for chronic pain (B**)**. Abbreviations: PTSD, post-traumatic stress disorder; OCD, obsessive–compulsive disorder; ADHD, attention-deficit/hyperactivity disorder; SAD, social anxiety disorder; PMDD, premenstrual dysphoric disorder.

**A)**
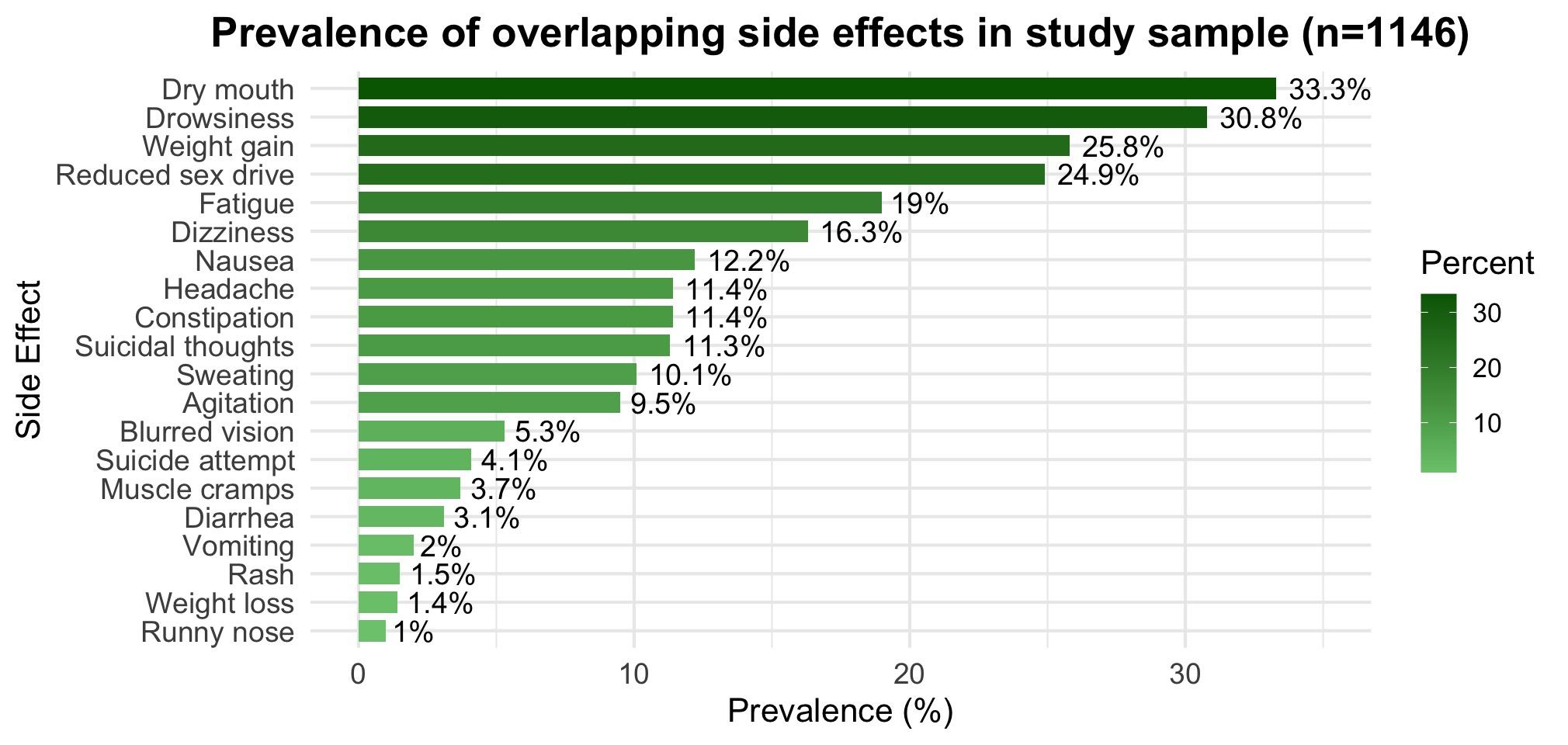

**B)**

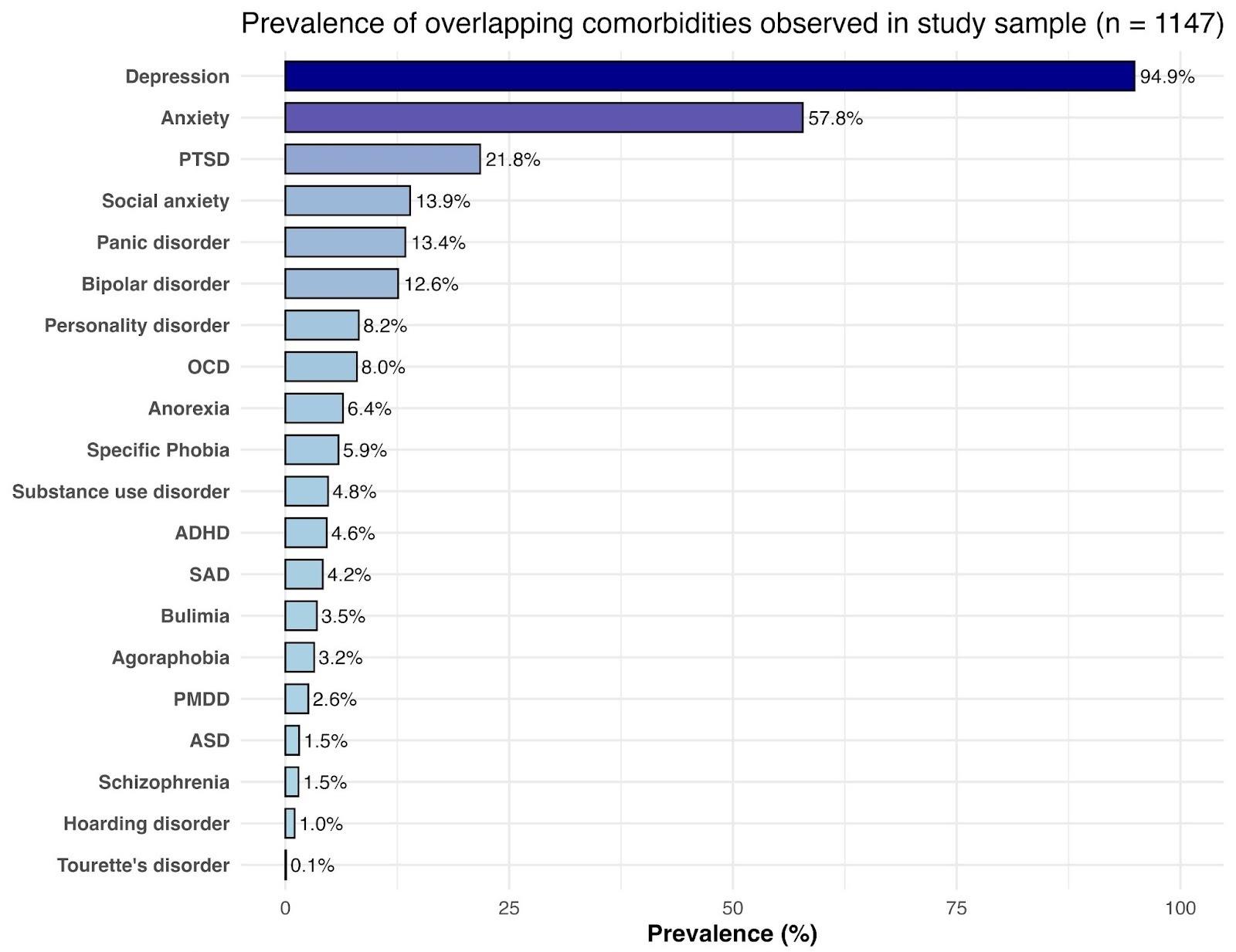

**Supplementary Figure 3.** Current (at the time the question was asked) and non-time-sensitive average pain intensity.

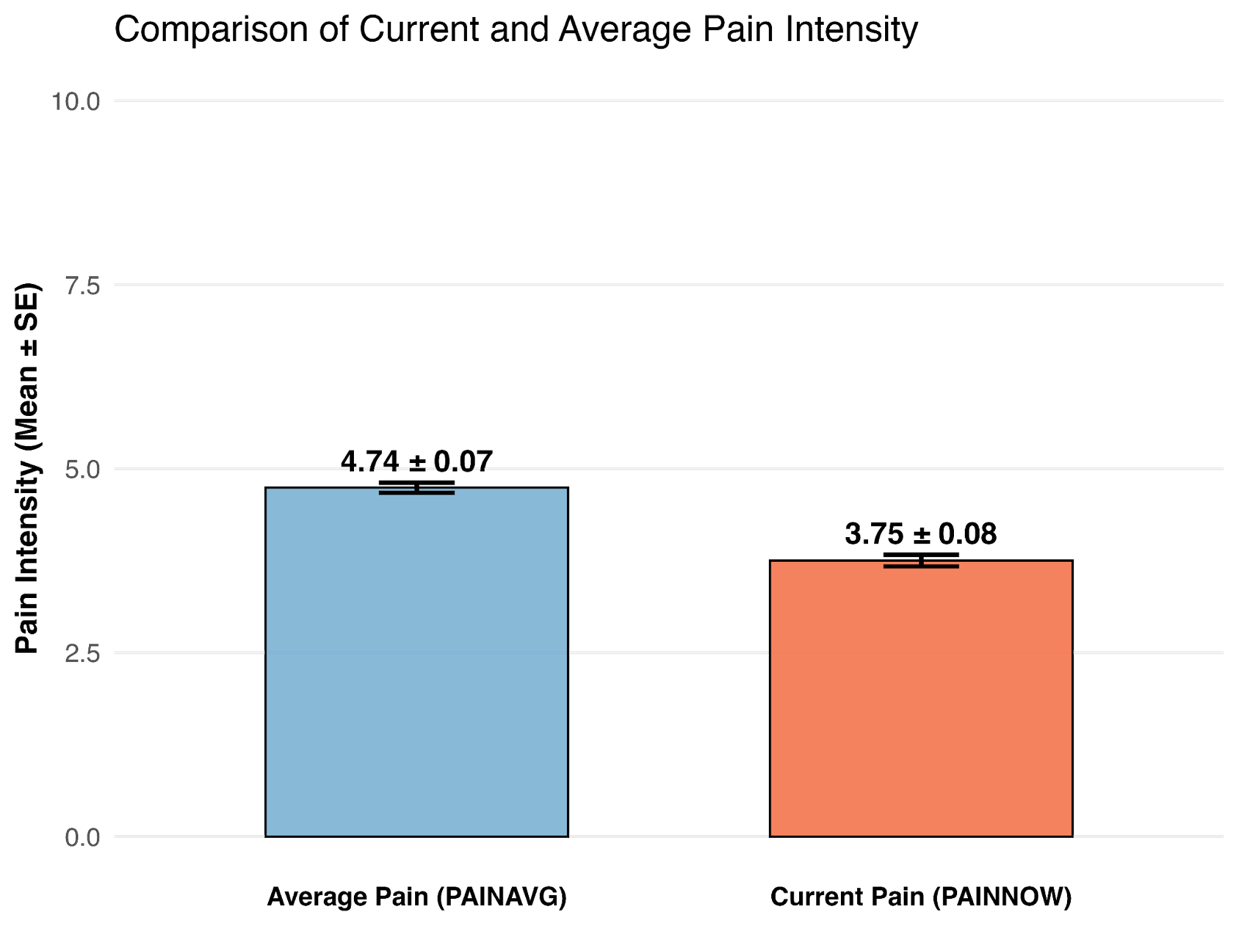
